# Longitudinal patterns of sensory and paranoid psychotic-like experiences in adolescents: Associations with distress, psychopathology, and substance use

**DOI:** 10.64898/2026.09.18.26363406

**Authors:** Frederic Berg, Grace Kiernan, Phuong Mi Nguyen, Maximilian Niggemeier, Silvia Schneider, Mar Rus-Calafell

## Abstract

**Background:** Psychotic-like experiences (PLEs) have been shown to increase the risk for mental health problems in later life, especially psychosis. Following previously raised methodological concerns about the assessment of PLEs in adolescents, this study sought to differentiate subtypes of PLEs and to explore their trajectories.

**Methods:** Longitudinally matched data of 671 German adolescents from three surveys over the time span of one year were analysed. Four trajectories were identified, including persisting, subsiding, and emerging highly indicative PLEs (HIPs), and non-highly indicative PLEs (non-HIPs), and longitudinally compared in terms of associated distress, psychopathology, and substance use. A differentiated never PLEs group was considered as a control.

**Results:** The persisting HIPs group reported greater distress than the non-HIP group over time and was consistently associated with higher rates of internalising symptoms. The subsiding HIP group showed significant reductions in distress and internalising symptoms over time, but no significant changes regarding externalising symptoms and substance use. There was no difference in the extent of substance use between the groups.

**Conclusions:** Screenings of PLEs should be mindful of different subtypes of PLEs that indicate a greater associated risk of poor mental health. PLE screening should not stop at whether PLEs are present but ask which ones.

## 1. Introduction

Psychotic-like experiences (PLE) are typically defined as subclinical psychotic symptoms occurring in the absence of illness (Kelleher & Cannon, 2011). Prevalence estimates vary, but approximately 7.5% of adolescents report them (Healy et al., 2019; Kelleher et al., 2012). While PLEs may be non-distressing, distressing PLEs appear to confer greater risk. Indeed, higher distress associated with attenuated psychotic symptoms predicts poorer clinical trajectories and non-remission of Clinical High Risk of Psychosis (CHRp; Nelson et al., 2022; Yung et al., 2005), suggesting that distressing PLEs constitute a particularly salient risk factor for psychotic disorders. However, more research exploring this is needed, since effective interventions for CHRp populations are lacking (Minichino et al., 2025).

Beyond psychosis risk, PLEs are associated with broader burdening psychopathology: they predict later depression and anxiety and are associated with elevated internalising symptoms in youth (Isaksson et al., 2020; Stainton et al., 2021). Certain PLE subtypes, such as ideas of reference and unusual perceptual experiences, appear particularly linked to anxiety (Nguyen et al., 2026; Unterrassner et al., 2017), and persistent or emerging PLEs are associated with increases in anxiety and depressive symptoms over time (Yamasaki et al., 2018; Xi et al., 2026). In contrast, associations with externalising symptoms are less consistent. Isaksson et al. (2020) found that baseline PLEs did not predict externalising symptoms at follow-up, although they were linked to later behavioural problems. Other studies suggest a reverse temporal relationship, with childhood externalising behaviours predicting adolescent PLEs (Lancefield et al., 2016; Wong et al., 2021). Together, these findings suggest that internalising symptoms may both precede and follow PLEs, whereas externalising behaviours may act primarily as antecedents.

Several risk factors have been implicated in the occurrence and persistence of PLEs, most notably substance use. Cannabis use is associated with increased psychosis risk in a dose-dependent manner, with frequent use (weekly or daily) conferring greater risk than rare or no use (Hasan et al., 2020; Robinson et al., 2023). Cannabis use may strengthen the association between PLEs and later psychotic disorder (Bourque et al., 2017; Denissoff et al., 2022). Cigarette smoking has also been consistently associated with PLEs across adolescent and adult samples (Barkhuizen et al., 2019; Mallet et al., 2018; Saha et al., 2011) and may predict psychosis onset (Kendler et al., 2015).

Findings across studies on various risk factors predicting incidence and persistence of PLEs remain inconsistent, and replication is limited (Staines et al., 2023c). The interpretation of associations is complicated by heterogeneity in how PLEs are conceptualised and measured. Studies vary in terminology, included phenomena, and assessment methods (Hinterbuchinger & Mossaheb, 2021; Lee et al., 2016), and PLE subtypes show differing associations with distress and psychopathology (Unterrassner et al., 2017; Yung et al., 2009), so reliance on composite PLE scores may mask clinically relevant risk patterns. Interpretation is further complicated by discrepancies between self-report and interview-based assessments: self-report measures may overestimate PLE prevalence, particularly for delusional ideation (Nelson et al., 2012), whereas interviews may underestimate PLEs due to non-disclosure (Gundersen et al., 2019). Agreement between methods is strongest for hallucinations and delusions, particularly paranoid delusions (Gundersen et al., 2019; Kelleher et al., 2011; Sun et al., 2022). Accordingly, recent reviews have restricted PLE definitions to subclinical hallucinations and delusions, which may better capture true positives and identify the PLEs most strongly associated with adverse mental health outcomes (Healy et al., 2019; Staines et al., 2023c).

The present study aimed to examine whether specific PLE subtypes, and their emergence or remission over time, are differentially associated with known risk factors. Building on previously established group classification (Kiernan et al., 2025], highly indicative and non-highly indicative PLEs were distinguished to identify distinct developmental trajectories differing in distress, psychopathology, and substance use. Following endorsement of experiences across time points, participants were classified into four PLE trajectory groups and one control group to examine both between-group differences and symptom-related changes over time: *Persisting highly indicative PLEs (HIP) Group*; non-highly indicative PLEs (non-HIP) *Group*; *Onset Group*; *Subsiding Group*; and *Never PLEs Group* (control group).

We hypothesised that 1) compared with non-highly indicative PLEs (*non-HIP Group*), persisting highly indicative PLEs (*Persisting HIP Group)* would be associated with higher PLE-associated distress, elevated anxiety, depression, and externalising symptoms, and greater substance use (tobacco and cannabis) over time; 2) individuals reporting exclusively non-highly indicative PLEs (*non-HIP Group)* were expected to show lower and more stable levels of distress, psychopathology and substance use over time; 3) the emergence of highly indicative PLEs (*Onset Group)* was hypothesised to coincide with increases in distress, psychopathology, and substance use over time; 4) the subsidence of highly indicative PLEs (*Subsiding Group)* would be associated with corresponding reductions in distress, psychopathology, and substance use over time.

## 2. Methods

The present study follows a prospective, longitudinal cohort design, consisting of a baseline data collection at the end of 2021 (BL) and two follow-ups (FU6 and FU12). The study was preregistered on OSF (Berg & Rus-Calafell, 2002) and approved by the local Ethics Committee of the Faculty of Psychology at the Ruhr-Universität Bochum (632/R1).

### 2.1 Participants

Participating adolescents were aged 12 to 19 years in North Rhine-Westphalia (Nguyen et al., 2026). Eligible participants were students at cooperating schools, with sufficient proficiency in German to comprehend the research procedures and the assessment booklet, and the capacity to provide informed consent. For participants under 16, parents were informed about the study and allowed to opt their children out. All participants had the right to withdraw from the study at any time and for any reason. Assessments were conducted at the educational centres at baseline, at 6-month (FU6), at 12-month (FU12), and at 24-month follow-up (FU24). There were 1,184 complete cases at baseline, 1164 at FU6, and 1,063 at FU12. Analyses included only participants with complete data at baseline, FU6, and FU12 (N=671).

### 2.2 Measures

Participants were asked to provide information about age, gender, first language, and country of citizenship. Socioeconomic status (SES) was assessed using the German version of the Family Affluence Scale (FAS) (Currie et al., 1997; Richter & Klocke, 2005).

The Psychotic Experiences Inventory (PEI) is an 11-item self-report questionnaire (Kelleher, 2024) to assess psychotic experiences, including perceptual and ideational disturbances, diminished emotional expression, and avolition. With permission from the authors, the questionnaire was translated into German and back-translated by an independent editor. Final approval for the translated version was obtained from the original developers. The PEI demonstrated excellent internal consistency (Cronbach’s α = .91) and very good convergent validity with the Community Assessment of Psychic Experiences 15 (*r* = .69, *p* < .001) (Mossaheb et al., 2012). The sensitivity and positive-predictive value (PPV) of the items vary widely, with the items assessing auditory or visual hallucinations and paranoid thoughts still showing the best predictive power (Kelleher et al., 2011). Following group classification in a subsample of this cohort (Kiernan et al., 2025), these three items were considered highly indicative PLEs (HIPs), while the other eight were considered non-highly indicative PLEs (non-HIPs). PLE-associated distress was calculated by dividing the sum of distress that adolescents reported by the individual number of PLEs present.

The Revised Children’s Anxiety and Depression Scale – Short Version (RCADS-25) is a self-report measure designed to assess symptoms of anxiety and depression in children and adolescents (Ebesutani et al., 2012). Items are rated on a 4-point Likert scale ranging from 0 (never) to 3 (always). In the present study, internal consistency at baseline was good for both the anxiety and depression subscales, with Cronbach’s α = .85 for anxiety (FU6, α = .86; FU12, α = .87) and α = .89 for depression (FU6, α = .89; FU12, α = .89).

The Strengths and Difficulties Questionnaire (SDQ) is a self-report measure consisting of five subscales (Goodman et al., 1998): emotional symptoms, conduct problems, hyperactivity/inattention, peer relationship problems, and prosocial behaviour. Items are rated on a 3-point Likert scale from 0 (not true) to 2 (certainly true). Here, only the externalising problems scale was included in the analyses. Internal consistency for this scale was questionable to acceptable, with Cronbach’s α = .68 at baseline and α = .72 at FU12; the scale was not administered at FU6.

Substance use (tobacco and cannabis) was assessed using an adapted version of the HBSC survey (Roberts et al., 2007) as adapted and used by Frobel et al. (2022). Tobacco use was measured with the item “How often do you smoke?” rated on a 4-point scale (0 = I do not smoke or have quit to 3 = I smoke every day). Cannabis use was assessed by frequency of use in the past six months (0 = never to 6 = 40 or more times or days), as well as age at first use, reported in months or years. For analysis, cannabis response categories were converted to estimated numbers of use (occasions/days) using category midpoints (e.g., 3–5 times/days = 4), whereas tobacco use was retained on its original ordinal scale. Substance use measures were collected at FU6 and FU12.

### 2.3 Procedure

A total of 205 secondary education centres of North Rhine-Westphalia (Germany) were contacted by post or email to participate in the main longitudinal study (Nguyen et al., 2026). Seven centres agreed to collaborate and allowed data collection in their centres. Students and parents were informed 2 weeks before every data collection period about the study. On data collection dates, students were asked to sign a consent form and were given an assessment booklet to complete. Each participating class was supervised and instructed on how to complete the assessment booklet by a research study member.

### 2.4 Statistical Analysis

All statistical analyses were conducted in R for Windows (version 4.3.2; R Core Team, 2020). Descriptive statistics are reported for the final sample (Table 1). All hypothesis tests were two-tailed with a significance threshold of *p* < .05. Missing data were handled using complete-case analyses; no imputation was performed. Given the study’s focus on associations between PLEs and psychopathology, potential outliers were retained.

**Table 1.** Baseline Demographics for Total Sample and PLE Trajectory Groups.

|  | Total Sample<br>*N* = 671 | Never PLEs<br>*n* = 44<br>(6.56%) | Persisting HIP<br>*n* = 279<br>(41.58%) | Non-HIP<br>*n* = 117<br>(17.44%) | Onset<br>*n* = 100<br>(14.91%) | Subsiding<br>*n* = 99<br>(14.75%) | Group<br>Comparisons<br>*p*-values<sup>a</sup> | Excluded<br>*n* = 32<br>(4.76%) |
| --- | --- | --- | --- | --- | --- | --- | --- | --- |
| Age (*M*, *SD*)<sup>b</sup> | 14.65 (1.01) | 14.18 (0.90) | 14.69 (0.98) | 14.63 (0.97) | 14.58 (0.98) | 14.88 (1.11) | .003* | 14.50 (1.08) |
| Gender |  |  |  |  |  |  | <.001*† |  |
| Female | 339 (50.52%) | 18 (40.91%) | 166 (59.50%) | 51 (43.59%) | 43 (43.00%) | 46 (46.46%) |  | 15 (46.88%) |
| Male | 318 (47.39%) | 26 (59.09%) | 103 (36.92%) | 65 (55.56%) | 57 (57.00%) | 50 (50.51%) |  | 17 (53.12%) |
| Diverse | 10 (1.49%) | 0 (0.00%) | 8 (2.87%) | 0 (0.00%) | 0 (0.00%) | 2 (2.02%) |  | 0 (0.00%) |
| Missing Info | 4 (0.60%) | 0 (0.00%) | 2 (0.72%) | 1 (0.85%) | 0 (0.00%) | 1 (1.01%) |  | 0 (0.00%) |
| Grade at Baseline |  |  |  |  |  |  | .011* |  |
| Grade 8 | 239 (35.62%) | 24 (54.55%) | 86 (30.82%) | 50 (42.74%) | 33 (33.00%) | 29 (29.29%) |  | 17 (53.12%) |
| Grade 9 | 267 (39.79%) | 17 (38.64%) | 118 (42.29%) | 41 (35.04%) | 45 (45.00%) | 42 (42.42%) |  | 4 (12.50%) |
| Grade 10 | 163 (24.29%) | 2 (4.55%) | 74 (26.52%) | 26 (22.22%) | 22 (22.00%) | 28 (28.28%) |  | 11 (34.38%) |
| Missing Info | 2 (0.30%) | 1 (2.27%) | 1 (0.36%) | 0 (0.00%) | 0 (0.00%) | 0 (0.00%) |  | 0 (0.00%) |
| First Language |  |  |  |  |  |  | .151† |  |
| German | 502 (74.81%) | 38 (86.36%) | 193 (69.18%) | 93 (79.49%) | 80 (80.00%) | 72 (72.73%) |  | 26 (81.25%) |
| English | 1 (0.15%) | 0 (0.00%) | 0 (0.00%) | 0 (0.00%) | 1 (1.00%) | 0 (0.00%) |  | 0 (0.00%) |
| French | 4 (0.60%) | 1 (2.27%) | 2 (0.72%) | 0 (0.00%) | 0 (0.00%) | 1 (1.01%) |  | 0 (0.00%) |
| Polish | 7 (1.04%) | 0 (0.00%) | 3 (1.08%) | 3 (2.56%) | 0 (0.00%) | 0 (0.00%) |  | 1 (3.12%) |
| Russian | 8 (1.19%) | 0 (0.00%) | 5 (1.79%) | 0 (0.00%) | 1 (1.00%) | 1 (1.01%) |  | 1 (3.12%) |
| Turkish | 30 (4.47%) | 2 (4.55%) | 13 (4.66%) | 2 (1.71%) | 3 (3.00%) | 8 (8.08%) |  | 2 (6.25%) |
| Other | 49 (7.30%) | 2 (4.55%) | 29 (10.39%) | 8 (6.84%) | 6 (6.00%) | 3 (3.03%) |  | 1 (3.12%) |
| Missing Info | 70 (10.43%) | 1 (2.27%) | 34 (12.19%) | 11 (9.40%) | 9 (9.00%) | 14 (14.14%) |  | 1 (3.12%) |
| Socioeconomic Status |  |  |  |  |  |  | .454† |  |
| Low | 9 (1.34%) | 0 (0.00%) | 8 (2.87%) | 0 (0.00%) | 1 (1.00%) | 0 (0.00%) |  | 0 (0.00%) |
| Medium | 112 (16.69%) | 8 (18.18%) | 52 (18.64%) | 19 (16.24%) | 14 (14.00%) | 15 (15.15%) |  | 4 (12.50%) |
| High | 550 (81.97%) | 36 (81.82%) | 219 (78.49%) | 98 (83.76%) | 85 (85.00%) | 84 (84.85%) |  | 28 (87.50%) |

*Note.* a PLE = psychotic-like experience; HIP = highly indicative PLE; non-HIP = non-highly indicative PLE If not specified otherwise, groups were compared by conducting non-parametric Kruskal-Wallis tests for numerical or $\chi^2$ -tests for categorical variables. Fisher-Exact-Tests in cases of empty cells are indicated by '†'. Missing values were not considered in group comparisons. *Asterisks* indicate significant values. b *M* = Mean, (*SD*) = Standard Deviation. c Subsequent values presented are *n* = Number of Cases, (%) = Percentage Proportion

#### 2.4.1 Group criteria and analysis of group differences

Participants were classified into four PLE trajectory groups and one control group using predefined criteria. The Persisting HIP group included individuals reporting at least one highly indicative PLE at baseline (BL), FU6, and FU12, or at BL and FU12 only. The non-HIP group comprised individuals reporting at least one non-highly indicative PLE at any assessment but no highly indicative PLEs at any time. The Onset group consisted of individuals who did not report HIPs at BL but did so at FU12 or at both FU6 and FU12. The Subsiding group included individuals who reported HIPs at BL or at BL and FU6, but no longer at FU12. Participants not reporting any PLEs at any assessment were assigned to the Never PLEs control group. Individuals not meeting criteria for any group were excluded from trajectory analyses. Group differences in demographic characteristics at baseline were examined using Kruskal–Wallis tests for continuous variables and χ² tests for categorical variables.

#### 2.4.2 PLE trajectory analysis

PLE-associated distress, depression, anxiety, and externalising problems were analysed using two generalised linear mixed-effects models (GLMMs) with random intercepts and random slopes for time for each outcome. Model 1 included Time of Measurement, PLE Trajectory Group, and their interaction as fixed effects, with gender, SES, and age entered as covariates. The reference group was the non-HIP group.

Interactions between Time of Measurement and PLE-Trajectory group were examined to assess whether the emergence or subsidence of HIPs was associated with differential change over time relative to the non-HIP group. The results from the respective Model-1 highlight differences in the groups’ slopes. Since the Onset and Subsiding groups are inherently associated with a change in the overall amount of reported PLEs, Time × Group interactions were only interpreted based on the respective Model 1.

Model 2 additionally adjusted for the total number of reported PLEs to distinguish effects attributable to PLE subtype beyond overall symptom burden. For depression, anxiety, and externalising problems, PLE-associated distress was included as an additional covariate to examine whether group differences in general psychopathology were explained by elevated distress associated with HIPs.

Cannabis use at FU6 and FU12 was analyzed using a GLMM, with the same fixed- and random-effects structure as Model 1. Tobacco use was initially analysed using mixed ordinal logistic regression with the same fixed- and random-effects structure. When this model failed to converge, tobacco use was analysed as a binary outcome (current smoker vs. non-smoker) using a binomial GLMM with a logit link function.

Relevant effect sizes (Cohen’s *d*) were calculated for significant between- and within-group differences, derived from paired comparison estimates and pooled standard deviations (see supplementary materials S2).

## 3. Results

### 3.1 Sample characteristics and trajectory group allocation

Matching data across all three assessment points resulted in a final sample of *N* = 671 adolescents. Baseline characteristics are summarised in Table 1. The prevalence of HIPs varied substantially across subtypes and time points. Auditory hallucinations were the least likely, with 13.71% at BL, 11.62% at FU6 and 10.43% at FU12, followed by visual hallucinations with 19.23% at BL, 17.73% at FU6 and 19.08% at FU12. In comparison, self-reported paranoid ideation was far more common with 49.48% at BL, 47.24% at FU6 and 51.27% at FU12.

Figure 1 depicts the mean number of self-reported PLEs across trajectory groups (See also S1). At baseline, the median number of PLEs was low in the non-HIP group and substantially higher in the Persisting HIP group. This difference remained evident at FU6 and FU12, despite considerable within-group variability. As expected, the number of PLEs increased over time in the Onset group and decreased in the Subsiding group, supporting the validity of the trajectory classification. Supplementary materials (S2 and S3) provide a full overview of model estimates.

**Figure 1.**
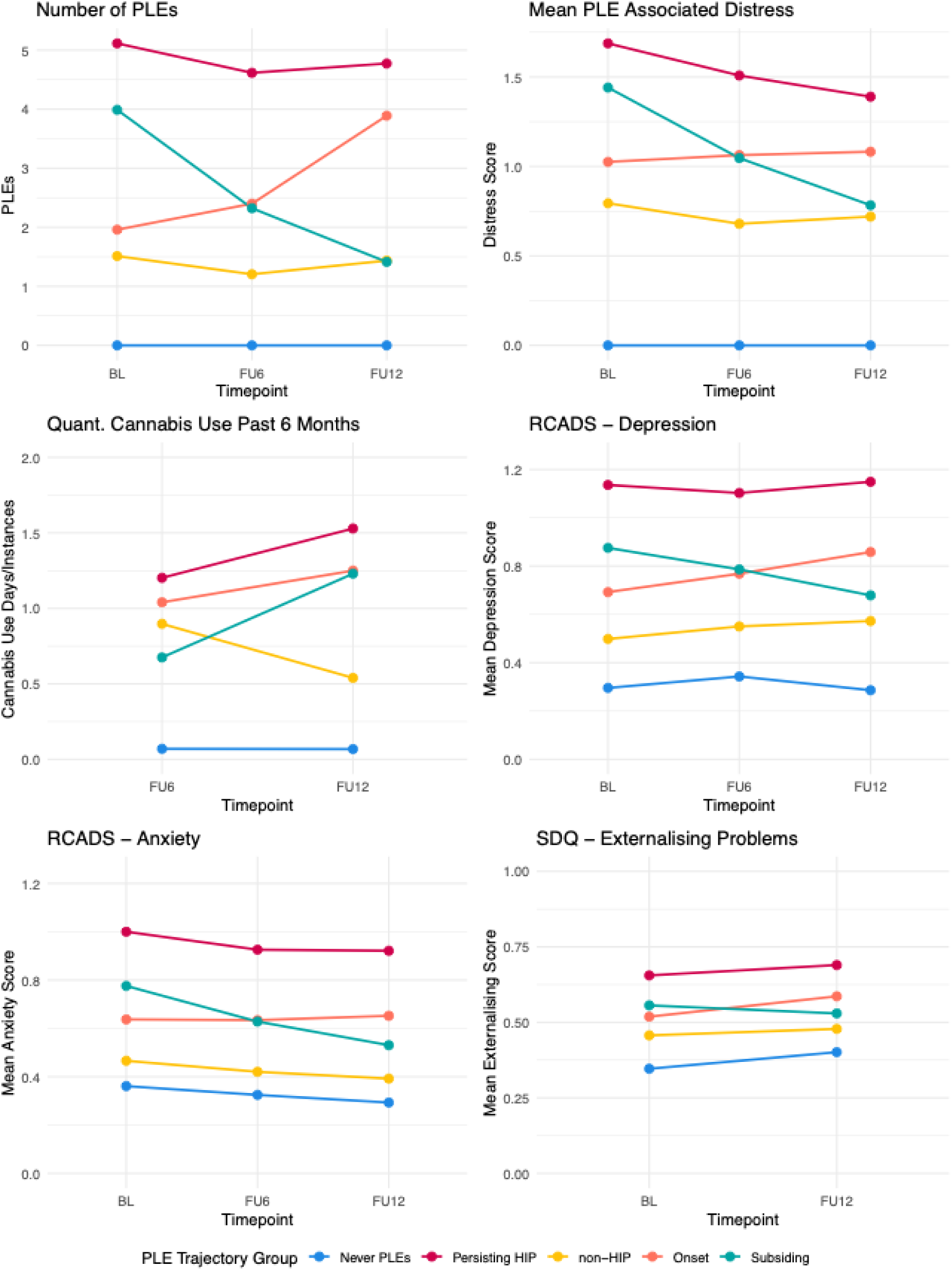
Overview of PLE-Trajectory Group Differences. *Note:* Plots depict group trajectories regarding the number of PLEs and associated distress, cannabis and tobacco use, and depression, anxiety, and externalising symptoms. Please note that the depicted plots are only a visual aid to the trajectory analysis further detailed in the Results section. Ordinates do not depict the whole range of the presented dependent variable to better visualise group differences.

At baseline, significant group differences were observed for age, gender, and grade (Table 1). Post hoc tests indicated that adolescents in the Never PLEs group were significantly younger than those in the other four groups, with no age differences among the remaining groups. This pattern was mirrored for grade, with a higher proportion of Grade 8 students in the Never PLEs group.

### 3.2 Persisting HIP vs. non-HIP (Hypothesis 1 & 2)

To test Hypotheses 1 and 2, differences between individuals with persisting HIPs and those reporting exclusively non-HIPs were examined using two sequential models.

In Model 1, individuals with persisting HIPs had substantially higher levels of PLE-associated distress, depression, anxiety, and externalising symptoms (S2) than the non-HIP group over time (all *p* < .001). Group differences were large for distress, depression, and anxiety, and medium-sized for externalising symptoms (S2).

In Model 2, additionally controlling for the total number of reported PLEs, group differences were attenuated but remained statistically significant for distress and internalising symptoms (all *p ≤* .004) (S2). Group differences did not remain significant for externalising symptoms (*p* = .578). This pattern indicates that elevated psychopathology was not solely attributable to a greater overall PLE burden, but also to the presence of HIPs. Furthermore, in line with Hypothesis 2, individuals reporting exclusively non-HIPs therefore showed lower and relatively stable levels of distress and general psychopathology over time. Neither cannabis nor tobacco use differed significantly between the Persisting HIP and non-HIP groups, and there was no evidence of differential change over time (all *p* > .05 (S3).

### 3.3 Emergence of HIPs (Hypothesis 3)

For the Onset group, there were no significant differences in trajectories of distress, depression, anxiety, externalising symptoms, or substance use compared with the non-HIP group (all *p* > .05), indicating that the emergence of HIPs was not accompanied by significant parallel changes in psychopathology or substance use during the observation period.

### 3.4 Subsidence of HIPs (Hypothesis 4)

Partially consistent with Hypothesis 4, the subsidence of HIPs was associated with significant reductions in PLE-associated distress and internalising symptoms (all *p ≤* .002) over time. The Subsiding group showed a significantly steeper decline in PLE-associated distress (*p* < .001), depressive symptoms (*p* = .001) and anxiety (*p* = .002) compared with the non-HIP group at FU12 (S2). These findings indicate that reductions in HIPs were accompanied by meaningful improvements in distress and internalising symptoms. In contrast, changes in externalising symptoms did not significantly differ between the Subsiding and non-HIP groups (*p* = .285) (S2), nor did changes in cannabis or tobacco use (*p* = .188 and *p* = .439, respectively) (S3).

## 4. Discussion

This study examined whether PLE subtypes and trajectories were differentially associated with distress, psychopathology, and substance use. Persistent HIPs were associated with greater distress, depression, and anxiety than non-HIPs, even after accounting for total PLE burden. In contrast, substance use and externalising symptoms did not distinguish the groups.

Consistent with previous studies (Unterrassner et al., 2017; Yung et al., 2009), PLE subtypes were differentially associated with distress and psychopathology. This is consistent with network-analytic findings from the same cohort, where positive PLEs were most strongly and consistently associated with anxiety across three assessment waves, while negative PLEs were most strongly linked to depression (Nguyen et al., 2026). While most individuals only experience transitory PLEs (Staines et al., 2023c), and these are still associated with adverse outcomes (Staines et al., 2023b), persisting PLEs seem to be especially concerning (e.g., Rimvall et al., 2020; Yamasaki et al., 2018), particularly when they involve highly-indicative phenomena such as hallucinations and delusions (HIPs): In this study, individuals reporting persisting HIPs exhibited consistently higher rates of internalising symptoms, including anxiety, than those experiencing only non-HIPs, mirroring the strong and stable positive PLE–anxiety association identified in the same cohort using network analysis (Nguyen et al., 2026). This is further supported by experimental evidence from a subsample of this cohort, where adolescents in the HIP group reported significantly higher social anxiety, avoidance, and fear of negative evaluation than the non-HIP group during real-time virtual social interactions, despite no corresponding differences in physiological arousal (Kiernan et al., 2025). Non-HIPs may be unable to discriminate from internalising symptoms, since, for example, “lack of motivation” can reflect both a non-HIP and a symptom of depression; this would be in line with difficulties distinguishing depressive features from negative symptoms in psychosis (Krynicki et al., 2018).

Unlike Yamasaki et al. (2018), who found that a remitting PLE trajectory was not accompanied by significant reductions in depression and anxiety, subsiding HIPs in this study were associated with a decrease in internalising symptoms over time. Furthermore, the subsidence of HIPs over time was also accompanied by a reduction in distress, suggesting that the resolution of HIPs is what may drive improvement in distress. However, without subsequent interview-based verification of these experiences, and given the self-reported/interview discrepancies discussed above, there remains greater uncertainty about whether these non-HIPs constituted true PLEs. Both persistence as well as PLE subtype therefore appear relevant to PLE-associated distress and to associated psychopathology.

While subsidence of HIPs was significantly associated with a reduction of psychopathology, there was no difference in psychopathology trajectories between the Onset and the non-HIP group. One possible explanation for the subsidence finding is that resolution of HIPs quickly alleviated co-occurring symptoms; alternatively, the reverse causal pathway may hold, whereby decreasing depression or anxiety contributes to the alleviation of HIPs. The absence of a corresponding effect for onset, meanwhile, may reflect a delay between the emergence of HIPs and their psychological impact: the onset of HIPs might not immediately aggravate other mental health issues, as individuals may need time to make sense of these experiences and subsequently appraise them as distressing. This would be consistent with cognitive models of psychosis (Garety et al., 2001), which emphasize the role of appraisal in the development and persistence of psychotic experiences. Associated psychopathology might not arise until these experiences persist over prolonged periods of time (Xi et al., 2026; Yamasaki et al., 2018).

Externalising symptoms did not differ between the groups. While the association of, for example, anxiety and PLEs seems intuitive, since the unusual experiences might frighten the individual, externalising behaviour might not be a logical consequence of PLEs. Conversely, previous research described externalising problems as antecedent to emerging PLEs (Isaksson et al., 2020; Lancefield et al., 2016), a finding this study could not support.

Trajectories of substance use also did not differ between the PLE trajectory groups, contrasting with previous research linking cannabis and tobacco use to PLEs (Barkhuizen et al., 2019; Bourque et al., 2017; Matheson et al., 2022) and with findings of bidirectional associations between PLEs and substance abuse (Degenhardt, 2018). Given that substance use was assessed only at FU6 and FU12, further longitudinal research is needed to clarify the temporal relationship between substance use and PLEs in adolescence.

Several limitations should be considered when interpreting these findings. First, PLEs were assessed exclusively via self-report using a brief 11-item screening tool, without subsequent interview-based verification; as such, the possibility of overestimated prevalence, particularly for delusional ideation, cannot be excluded (Nelson et al., 2012), and item-level distinctions between HIPs and non-HIPs, while grounded in prior classification work in a subsample of this cohort (Kiernan et al., 2025), would benefit from replication using more detailed or interview-based PLE measures. Second, substance use was assessed only at FU6 and FU12, precluding examination of baseline associations and limiting inference about temporal ordering between PLEs and substance use; self-reported cannabis and tobacco use may also be subject to underreporting given that both substances are illegal for minors in Germany. Third, although the final analytic sample was reasonably large (N = 671), it represents a substantial reduction from the baseline sample (N = 1,184) due to the use of complete-case analysis without imputation, raising the possibility of selective attrition. Finally, the sample was drawn from secondary schools in a single German region and may not be representative of adolescents more broadly, particularly with respect to socioeconomic and school-type diversity; generalisability to more diverse or disadvantaged populations should therefore be made with caution.

Methodologically, the findings underscore prevailing issues in PLE research (Hinterbuchinger & Mossaheb, 2021) and suggest that PLE subtypes should be considered alongside self-reported measures; future research would benefit from a more universally accepted approach to defining and measuring PLEs. This study highlights PLEs as an important risk factor for poor mental health, with PLE subtypes providing relevant information beyond overall PLE number that may inform future risk prediction. Schools, parents, and health care providers should be aware of the relevance of PLEs and respond accordingly to young people who describe such experiences: school-based prevention programmes addressing PLEs have shown efficacy (Staines et al., 2023a), and parent-adolescent relationships appear to mediate the association between PLEs and adversity (McMahon et al., 2021), underscoring the central role both parents and health care providers can play if equipped with current knowledge on the issue. Preventing PLEs or alleviating associated distress might translate into fewer young people transitioning to more serious expressions of the psychosis spectrum. Future research should attend to different dimensions of PLEs and further investigate how substance use and the emergence of PLEs coincide over time, building on PLEs’ role as part of the psychosis continuum and their relevance to youth mental health prevention efforts.

## Data Availability

The data that support the findings of this study are not publicly available due to their sensitive nature but are available from the corresponding authors upon reasonable request. The data are stored in a controlled-access repository at the Department of Clinical Psychology and Digital Psychotherapy, Bochum, Germany.

## CRediT authorship contribution statement

**FB:** Conceptualization, Methodology, Investigation, Data curation, Formal analysis, Writing – original draft. **GK:** Investigation, Data curation, Writing – original draft, Writing – review & editing**. P-MN**: Investigation, Data curation. **MN**: Writing – review & editing. **SS**: Supervision. **MRC**: Funding acquisition, Project administration, Conceptualization, Methodology, Investigation, Data Curation, Supervision, Writing – review & editing.

## Acknowledgements

We would like to sincerely thank all schools including the teachers whose support was crucial in the data collection process. This study would not have been possible without the students who participated-thank you. Our thanks also go to the research assistants Fine Kullmann, Sandra Abrantes-Díaz, Max Braun Rodrigues, Pauline Kohl, Lisa Kahl, Dilara Alatas, and Maren Marzinzik for their dedication and assistance throughout the project. We would also like to thank Prof. Ian Kelleher (University of Edinburgh) for his invaluable advice and clinical guidance on assessment of PLEs in adolescent populations. We would also like to extend our warm thanks to Dr. Amy Hardy (King’s College London) for her invaluable guidance and support in adapting the TALE for the purposes of this study.

