## Supplementary material for "Longitudinal patterns of sensory and paranoid psychotic-like experiences in adolescents: Associations with distress, psychopathology, and substance use": Supplemtentary Material

### Supplementary Materials

**Table S1.** *Descriptive Statistics for Study Variables by PLE Trajectory Group and Time Point*

| Variable | PLE trajectory group | BL (M(SD)) | Median (Range) | FU6 (M(SD)) | Median (Range) | FU12 (M(SD)) | Median (Range) |
|---|---|---|---|---|---|---|---|
| Number of PLEs | Persisting HIP | 5.11 (2.47) | 5 (1–11) | 4.62 (2.63) | 4 (0–11) | 4.77 (2.33) | 4 (0–11) |
| Number of PLEs | non-HIP | 1.51 (1.42) | 1 (0–7) | 1.21 (1.30) | 1 (0–6) | 1.44 (1.35) | 1 (0–6) |
| Number of PLEs | Onset | 1.96 (1.52) | 2 (0–6) | 2.40 (1.91) | | 3.89 (1.98) | 4 (1–11) |
| Number of PLEs | Subsiding | 3.99 (2.07) | 4 (1–10) | 2.32 (2.11) | | 1.41 (1.45) | 1 (0–6) |
| PLE-Associated Distress | Never PLEs | 0.00 (0.00) | | 0.00 (0.00) | | 0.00 (0.00) | |
| PLE-Associated Distress | Persisting HIP | 1.69 (0.75) | | 1.51 (0.83) | | 1.39 (0.76) | |
| PLE-Associated Distress | non-HIP | 0.79 (0.85) | | 0.68 (0.79) | | 0.72 (0.87) | |
| PLE-Associated Distress | Onset | 1.03 (0.90) | | 1.06 (0.88) | | 1.08 (0.76) | |
| PLE-Associated Distress | Subsiding | 1.44 (0.76) | | 1.05 (0.90) | | 0.78 (1.03) | |
| Cannabis Use Past 6 Months | Never PLEs | NA | | 0.07 (0.32) | | 0.07 (0.32) | |
| Cannabis Use Past 6 Months | Persisting HIP | NA | | 1.20 (5.32) | | 1.53 (6.04) | |
| Cannabis Use Past 6 Months | non-HIP | NA | | 0.90 (5.29) | | 0.54 (2.97) | |
| Cannabis Use Past 6 Months | Onset | NA | | 1.04 (5.76) | | 1.25 (6.38) | |
| Cannabis Use Past 6 Months | Subsiding | NA | | 0.68 (4.33) | | 1.23 (5.93) | |
| RCADS - Depression | Never PLEs | 0.30 (0.23) | | 0.34 (0.38) | | 0.29 (0.25) | |
| RCADS - Depression | Persisting HIP | 1.14 (0.60) | | 1.10 (0.56) | | 1.15 (0.57) | |
| RCADS - Depression | non-HIP | 0.50 (0.32) | | 0.55 (0.36) | | 0.57 (0.35) | |
| RCADS - Depression | Onset | 0.69 (0.45) | | 0.77 (0.52) | | 0.86 (0.47) | |
| RCADS - Depression | Subsiding | 0.88 (0.53) | | 0.79 (0.56) | | 0.68 (0.51) | |
| RCADS - Anxiety | Never PLEs | 0.36 (0.22) | | 0.32 (0.25) | | 0.29 (0.19) | |
| RCADS - Anxiety | Persisting HIP | 1.00 (0.47) | | 0.93 (0.48) | | 0.92 (0.48) | |
| RCADS - Anxiety | non-HIP | 0.47 (0.28) | | 0.42 (0.28) | | 0.39 (0.27) | |
| RCADS - Anxiety | Onset | 0.64 (0.33) | | 0.63 (0.37) | | 0.65 (0.37) | |
| RCADS - Anxiety | Subsiding | 0.78 (0.43) | | 0.63 (0.43) | | 0.53 (0.34) | |
| SDQ - Externalising | Never PLEs | 0.35 (0.28) | | NA | | 0.40 (0.34) | |
| SDQ - Externalising | Persisting HIP | 0.66 (0.31) | | NA | | 0.69 (0.34) | |
| SDQ - Externalising | non-HIP | 0.46 (0.28) | | NA | | 0.48 (0.30) | |
| SDQ - Externalising | Onset | 0.52 (0.26) | | NA | | 0.59 (0.33) | |
| SDQ - Externalising | Subsiding | 0.56 (0.36) | | NA | | 0.53 (0.32) | |

*Note*. Values are presented as mean (*M*) and standard deviation (*SD*); medians and ranges are additionally reported for the number of PLEs. BL = baseline; FU6 = 6-month follow-up; FU12 = 12-month follow-up; PLE = psychotic-like experience; HIP = highly indicative psychotic-like experience; non-HIP = non-highly indicative psychotic-like experience; RCADS = Revised Children's Anxiety and Depression Scale; SDQ = Strengths and Difficulties Questionnaire; NA = not available.

**Table S2.** *PLE-Trajectory Analysis: Distress Models, Depression Models, Anxiety Models and Externalising Models*

| | Model 1 | | | | | | Model 2 | | | | | |
|---|---|---|---|---|---|---|---|---|---|---|---|---|
| | β | *SE* | *df* | *t* | *p* | *d* | β | *SE* | *df* | *t* | *p* | *d* |
| Distress-Models | | | | | | | | | | | | |
| Fixed Effects | | | | | | | | | | | | |
| (Intercept) | 1.119 | 0.217 | 1259 | 5.145 | <.001* | | 0.691 | 0.199 | 1258 | 3.467 | <.001* | |
| FU6 | -0.105 | 0.078 | 1259 | -1.342 | .180 | | -0.066 | 0.076 | 1258 | -0.874 | .382 | |
| FU12 | -0.062 | 0.083 | 1259 | -0.745 | .456 | | -0.053 | 0.081 | 1258 | -0.652 | .515 | |
| Group: Never PLEs | -0.764 | 0.134 | 626 | -5.696 | <.001* | | -0.570 | 0.126 | 626 | -4.528 | <.001* | |
| Group: Persisting HIP | 0.839 | 0.084 | 626 | 9.942 | <.001* | 1.09 (BL), 0.74 (FU12) | 0.405 | 0.084 | 626 | 4.817 | <.001* | 0.52 (BL), 0.25 (FU12) |
| Group: Onset | 0.236 | 0.103 | 626 | 2.287 | .023* | | 0.179 | 0.096 | 626 | 1.860 | .063 | |
| Group: Subsiding | 0.630 | 0.104 | 626 | 6.067 | <.001* | | 0.319 | 0.099 | 626 | 3.215 | .001* | |
| Gender Male | -0.298 | 0.049 | 626 | -6.023 | <.001* | | -0.197 | 0.045 | 626 | -4.374 | <.001* | |
| Gender Diverse | 0.052 | 0.195 | 626 | 0.268 | .789 | | -0.210 | 0.177 | 626 | -1.185 | .237 | |
| SES Medium | -0.208 | 0.212 | 626 | -0.983 | .326 | | 0.040 | 0.192 | 626 | -0.210 | .834 | |
| SES High | -0.150 | 0.206 | 626 | -0.727 | .468 | | 0.027 | 0.186 | 626 | 0.146 | .884 | |
| Age Scaled | 0.039 | 0.023 | 1259 | 1.715 | .087 | | 0.041 | 0.021 | 1258 | 1.923 | .055 | |
| Nr. of PLEs | – | – | – | – | – | | 0.129 | 0.009 | 1258 | 14.66 | <.001* | |
| Interactions | | | | | | | | | | | | |
| FU6 * Never PLEs | 0.105 | 0.149 | 1259 | 0.701 | .484 | | 0.066 | 0.144 | 1258 | 0.455 | .649 | |
| FU12 * Never PLEs | 0.063 | 0.158 | 1259 | 0.403 | .687 | | 0.054 | 0.154 | 1258 | 0.355 | .723 | |
| FU6 * Persisting HIP | -0.079 | 0.093 | 1259 | -0.850 | .396 | | -0.053 | 0.090 | 1258 | -0.584 | .559 | |
| FU12 * Persisting HIP | -0.246 | 0.098 | 1259 | -2.495 | .013* | | -0.209 | 0.096 | 1258 | -2.180 | .029* | |
| FU6 * Onset | 0.142 | 0.115 | 1259 | 1.232 | .218 | | 0.046 | 0.111 | 1258 | 0.412 | .680 | |
| FU12 * Onset | 0.116 | 0.121 | 1259 | 0.952 | .341 | | -0.143 | 0.120 | 1258 | -1.195 | .232 | |
| FU6 * Subsiding | -0.283 | 0.116 | 1259 | -2.446 | .015* | | -0.104 | 0.112 | 1258 | -0.930 | .353 | |
| FU12 * Subsiding | -0.594 | 0.122 | 1259 | -4.864 | <.001* | −0.71 | -0.269 | 0.121 | 1258 | -2.219 | .027* | |
| Random Effects | | | | | | | | | | | | |
| ID Var(SD) | | | 0.465 (0.682) | | | | | | 0.401 (0.633) | | | |
| FU6 Var(SD) | | | 0.502 (0.709) | | | | | | 0.471 (0.686) | | | |
| FU12 Var(SD) | | | 0.582 (0.763) | | | | | | 0.561 (0.749) | | | |
| Fit Indices | | | | | | | | | | | | |
| AIC | | | 4288.2 | | | | | | 4101.3 | | | |
| BIC | | | 4437.8 | | | | | | 4256.4 | | | |
| Depression-Models | | | | | | | | | | | | |
| Fixed Effects | | | | | | | | | | | | |
| (Intercept) | 0.885 | 0.144 | 1254 | 6.128 | <.001* | | 0.400 | 0.108 | 1252 | 3.690 | <.001* | |
| FU6 | 0.053 | 0.040 | 1254 | 1.311 | .190 | | 0.099 | 0.038 | 1252 | 2.630 | .009* | |
| FU12 | 0.077 | 0.045 | 1254 | 1.719 | .009* | | 0.096 | 0.038 | 1252 | 2.504 | .012* | |
| Group: Never PLEs | -0.201 | 0.085 | 626 | -2.357 | .019* | | 0.069 | 0.067 | 626 | 1.022 | .304 | |
| Group: Persisting HIP | 0.566 | 0.054 | 626 | 10.57 | <.001* | BL 1.06, FU12 0.97 | 0.130 | 0.045 | 626 | 2.919 | .004* | BL 0.24, FU12 0.25 |
| Group: Onset | 0.191 | 0.063 | 626 | 2.931 | .004* | | 0.113 | 0.051 | 626 | 2.218 | .027* | |
| Group: Subsiding | 0.350 | 0.066 | 626 | 5.320 | <.001* | | 0.033 | 0.053 | 626 | 0.627 | .531 | |
| Gender Male | 0.354 | 0.033 | 626 | 2.716 | .007* | | -0.167 | 0.025 | 626 | -6.756 | <.001* | |
| Gender Diverse | 0.354 | 0.130 | 626 | 2.716 | .007* | | 0.186 | 0.096 | 626 | 1.933 | .054 | |
| SES Medium | -0.250 | 0.142 | 626 | -1,764 | .078 | | -0.098 | 0.105 | 626 | -0.935 | .350 | |
| SES High | -0.222 | 0.137 | 626 | -1.612 | .108 | | -0.077 | 0.102 | 626 | -0.762 | .447 | |

| | Model 1 | | | | | | Model 2 | | | | | |
|---|---|---|---|---|---|---|---|---|---|---|---|---|
| | β | *SE* | *df* | *t* | *p* | *d* | β | *SE* | *df* | *t* | *p* | *d* |
| Age Scaled | 0.002 | 0.014 | 1254 | 0.150 | .881 | | -0.004 | 0.011 | 1252 | -0.330 | .741 | |
| Nr. of PLEs | – | – | – | – | – | | 0.084 | 0.005 | 1252 | 17.70 | <.001* | |
| PLE Distress | – | – | – | – | – | | 0.184 | 0.012 | 1252 | 15.85 | <.001* | |
| Interactions | | | | | | | | | | | | |
| FU6 * Never PLEs | 0.000 | 0.077 | 1254 | 0.004 | .997 | | -0.047 | 0.072 | 1252 | -0.653 | .514 | |
| FU12 * Never PLEs | -0.080 | 0.085 | 1254 | -0.938 | .348 | | -0.100 | 0.073 | 1252 | -1.376 | .169 | |
| FU6 * Persisting HIP | -0.083 | 0.048 | 1254 | -1.716 | .086 | | -0.052 | 0.045 | 1252 | -1.154 | .249 | |
| FU12 * Persisting HIP | -0.066 | 0.053 | 1254 | -1.252 | .211 | | 0.001 | 0.046 | 1252 | 0.012 | .991 | |
| FU6 * Onset | 0.023 | 0.059 | 1254 | 0.387 | .699 | | -0.067 | 0.056 | 1252 | -1.208 | .227 | |
| FU12 * Onset | 0.085 | 0.066 | 1254 | 1.289 | .198 | | -0.102 | 0.057 | 1252 | -1.797 | .073 | |
| FU6 * Subsiding | -0.139 | 0.060 | 1254 | -2.327 | .020* | | 0.028 | 0.056 | 1252 | 0.494 | .621 | |
| FU12 * Subsiding | -0.271 | 0.066 | 1254 | -4.123 | <.001* | −0.37 | 0.047 | 0.058 | 1252 | 0.824 | .410 | |
| Random Effects | | | | | | | | | | | | |
| ID Var(SD) | 0.194 (0.441) | | | | | | 0.114 (0.338) | | | | | |
| FU6 Var(SD) | 0.122 (0.349) | | | | | | 0.116 (0.341) | | | | | |
| FU12 Var(SD) | 0.162 (0.403) | | | | | | 0.120 (0.346) | | | | | |
| Fit Indices | | | | | | | | | | | | |
| AIC | 2154.4 | | | | | | 1540.5 | | | | | |
| BIC | 2303.9 | | | | | | 1701.1 | | | | | |
| Anxiety-Models | | | | | | | | | | | | |
| Fixed Effects | | | | | | | | | | | | |
| (Intercept) | 0.706 | 0.108 | 1254 | 6.541 | <.001* | | 0.388 | 0.090 | 1252 | 4.314 | <.001* | |
| FU6 | -0.044 | 0.032 | 1254 | -1.354 | .176 | | -0.013 | 0.031 | 1252 | -0.420 | .675 | |
| FU12 | -0.072 | 0.037 | 1254 | -1.926 | .054 | | -0.061 | 0.033 | 1252 | -1.838 | .066 | |
| Group: Never PLEs | -0.102 | 0.066 | 626 | -1.545 | .123 | | 0.068 | 0.057 | 626 | 1.201 | .230 | |
| Group: Persisting HIP | 0.466 | 0.041 | 626 | 11.28 | <.001* | BL 1.06, FU12 0.97 | 0.165 | 0.038 | 626 | 4.402 | <.001* | BL 0.24, FU12 0.25 |
| Group: Onset | 0.173 | 0.050 | 626 | 3.424 | <.001* | | 0.123 | 0.043 | 626 | 2.855 | .004* | |
| Group: Subsiding | 0.291 | 0.051 | 626 | 5.723 | <.001* | | 0.074 | 0.044 | 626 | 1.662 | .097 | |
| Gender Male | -0.307 | 0.025 | 626 | 5.723 | <.001* | | -0.233 | 0.020 | 626 | -11.40 | <.001* | |
| Gender Diverse | 0.221 | 0.097 | 626 | 2.278 | .023* | | 0.093 | 0.080 | 626 | 1.162 | .246 | |
| SES Medium | -0.062 | 0.105 | 626 | -0.587 | .558 | | 0.042 | 0.087 | 626 | 0.487 | .627 | |
| SES High | -0.068 | 0.102 | 626 | -0.661 | .509 | | 0.033 | 0.084 | 626 | 0.396 | .692 | |
| Age Scaled | -0.004 | 0.011 | 1254 | -0.405 | .685 | | -0.009 | 0.009 | 1252 | -0.969 | .333 | |
| Nr. of PLEs | – | – | – | – | – | | 0.067 | 0.004 | 1252 | 16.86 | <.001* | |
| PLE Distress | – | – | – | – | – | | 0.091 | 0.010 | 1252 | 9.414 | <.001* | |
| Interactions | | | | | | | | | | | | |
| FU6 * Never PLEs | 0.011 | 0.062 | 1254 | 0.175 | .861 | | -0.020 | 0.059 | 1252 | -0.335 | .738 | |
| FU12 * Never PLEs | 0.007 | 0.071 | 1254 | 0.096 | .923 | | -0.004 | 0.063 | 1252 | -0.070 | .944 | |
| FU6 * Persisting HIP | -0.028 | 0.038 | 1254 | -0.740 | .459 | | -0.008 | 0.037 | 1252 | -0.208 | .835 | |
| FU12 * Persisting HIP | -0.008 | 0.044 | 1254 | -0.178 | .859 | | 0.033 | 0.039 | 1252 | 0.830 | .407 | |
| FU6 * Onset | 0.041 | 0.047 | 1254 | 0.860 | .389 | | -0.023 | 0.045 | 1252 | -0.498 | .619 | |

| | Model 1 | | | | | | Model 2 | | | | | |
|---|---|---|---|---|---|---|---|---|---|---|---|---|
| | β | *SE* | *df* | *t* | *p* | *d* | β | *SE* | *df* | *t* | *p* | *d* |
| FU12 * Onset | 0.084 | 0.055 | 1254 | 1.533 | .126 | | -0.057 | 0.049 | 1252 | -1.172 | .242 | |
| FU6 * Subsiding | -0.103 | 0.047 | 1254 | -2.161 | .031* | | 0.015 | 0.046 | 1252 | 0.320 | .749 | |
| FU12 * Subsiding | -0.174 | 0.055 | 1254 | -3.184 | .002* | −0.63 | 0.047 | 0.050 | 1252 | 0.939 | .348 | |
| Random Effects | | | | | | | | | | | | |
| ID Var(SD) | 0.115 (0.339) | | | | | | 0.082 (0.286) | | | | | |
| FU6 Var(SD) | 0.077 (0.278) | | | | | | 0.077 (0.277) | | | | | |
| FU12 Var(SD) | 0.117 (0.342) | | | | | | 0.091 (0.302) | | | | | |
| Fit Indices | | | | | | | | | | | | |
| AIC | 1271.6 | | | | | | 852.7 | | | | | |
| BIC | 1421.2 | | | | | | 1013.3 | | | | | |
| Externalising-Models | | | | | | | | | | | | |
| Fixed Effects | | | | | | | | | | | | |
| (Intercept) | 0.600 | 0.099 | 625 | 6.053 | <.001* | | 0.426 | 0.092 | 625 | 4.644 | <.001* | |
| FU12 | 0.016 | 0.027 | 608 | 0.589 | .556 | | 0.022 | 0.027 | 606 | 0.832 | .406 | |
| Group: Never PLEs | -0.105 | 0.055 | 625 | -1.918 | .056 | | -0.005 | 0.051 | 625 | -0.094 | .925 | |
| Group: Persisting HIP | 0.187 | 0.034 | 625 | 5.513 | <.001* | BL 0.62, FU12 0.63 | 0.019 | 0.034 | 625 | 0.556 | .578 | |
| Group: Onset | 0.050 | 0.041 | 625 | 1.211 | .226 | | 0.021 | 0.038 | 625 | 0.548 | .584 | |
| Group: Subsiding | 0.092 | 0.042 | 625 | 2.201 | .028* | | -0.031 | 0.039 | 625 | -0.775 | .439 | |
| Gender Male | 0.025 | 0.023 | 625 | 1.071 | .284 | | 0.069 | 0.021 | 625 | 3.226 | .001* | |
| Gender Diverse | 0.253 | 0.090 | 625 | 2.809 | .005* | | 0.177 | 0.083 | 625 | 2.136 | .275 | |
| SES Medium | -0.146 | 0.098 | 625 | -1.494 | .136 | | -0.098 | 0.090 | 625 | -1.093 | .275 | |
| SES High | -0.152 | 0.095 | 625 | -1.600 | .110 | | -0.108 | 0.087 | 625 | -1.242 | .215 | |
| Age Scaled | -0.014 | 0.010 | 608 | -1.325 | 0.186 | | -0.018 | 0.010 | 606 | -1.929 | .054 | |
| Nr. of PLEs | – | – | – | – | – | | 0.034 | 0.004 | 606 | 8.025 | <.001* | |
| PLE Distress | – | – | – | – | – | | 0.066 | 0.010 | 606 | 6.323 | <.001* | |
| Interactions | | | | | | | | | | | | |
| FU12 * Never PLEs | 0.019 | 0.052 | 608 | 0.373 | .709 | | 0.014 | 0.051 | 606 | 0.276 | .783 | |
| FU12 * Persisting HIP | 0.020 | 0.032 | 608 | 0.625 | .532 | | 0.047 | 0.032 | 606 | 1.487 | .138 | |
| FU12 * Onset | 0.060 | 0.040 | 608 | 1.512 | .131 | | -0.012 | 0.040 | 606 | -0.298 | .766 | |
| FU12 * Subsiding | -0.043 | 0.040 | 608 | -1.069 | .285 | | 0.081 | 0.041 | 606 | 1.998 | .046* | |
| Random Effects | | | | | | | | | | | | |
| ID Var(SD) | 0.077 (0.278) | | | | | | 0.064 (0.252) | | | | | |
| FU12 Var(SD) | 0.056 (0.237) | | | | | | 0.056 (0.236) | | | | | |
| Fit Indices | | | | | | | | | | | | |
| AIC | 497.9 | | | | | | 383.5 | | | | | |
| BIC | 595.1 | | | | | | 490.9 | | | | | |

*Note.* Table S2 presents the results for the fixed and random effects of Distress-Models 1 and 2, Depression-Models 1 and 2, Anxiety-Models 1 and 2, Externalising-Models 1 and 2, as well as AIC and BIC for comparison of model fit. Baseline (BL)–ID is the random effect accounting for the random intercepts between individuals; The random effects for FU6 and FU12 are the variances and standard deviations for the random slopes over time. The interaction terms represent the moderation of the effect of time of measurement by each PLE trajectory group relative to the reference (non-HIP) group. Cohen's d values quantify selected significant between-group differences for the Persisting

HIP versus non-HIP groups and within-group BL-to-FU12 changes for the Subsiding group; SE = Standard Error; df = degrees of freedom; t = t-score for the corresponding estimate. Asterisks indicate statistical significance.

#### **Table S3.** *PLE-Trajectory Analysis: CannabisQuant Model and TobaccoBinary Model*

| | *CannabisQuant-Model* | | | | | *TobaccoBinary-Model* | | | |
|---|---|---|---|---|---|---|---|---|---|
| | β | *SE* | *df* | *t* | *p* | β | *SE* | *z* | *p* |
| **Fixed Effects** | | | | | | | | | |
| (Intercept) | 1.082 | 1.668 | 624 | 0.649 | .517 | -10.22 | 2.804 | 0.649 | <.001* |
| FU12 | -0.324 | 0.384 | 611 | -0.844 | .399 | 2.845 | 1.259 | 2.259 | .024* |
| Group: Never PLEs | -0.506 | 0.888 | 624 | -0.569 | .569 | 1.273 | 2.412 | 0.528 | .598 |
| Group: Persisting HIP | 0.311 | 0.560 | 624 | 0.554 | .580 | 0.762 | 1.606 | 0.474 | .635 |
| Group: Onset | 0.085 | 0.680 | 624 | 0.125 | .900 | 2.643 | 1.684 | 1.570 | .116 |
| Group: Subsiding | -0.339 | 0.688 | 624 | -0.493 | .622 | -2.020 | 2.178 | -0.927 | .354 |
| Gender Male | 0.621 | 0.387 | 624 | 1.604 | .109 | 0.488 | 0.735 | 0.664 | .506 |
| Gender Diverse | 0.724 | 1.520 | 624 | 0.477 | .634 | 0.724 | 1.520 | 0.477 | .634 |
| SES Medium | -1.486 | 1.653 | 624 | -0.899 | .369 | -1.662 | 2.484 | -0.669 | .503 |
| SES High | -0.293 | 1.604 | 624 | -0.183 | .855 | -1.041 | 2.299 | -0.453 | .651 |
| Age Scaled | 0.784 | 0.172 | 611 | 4.546 | <.001* | 0.622 | 0.361 | 1.723 | .085 |
| **Interactions** | | | | | | | | | |
| FU12*Never PLEs | 0.363 | 0.731 | 611 | 0.497 | .620 | -1.537 | 2.067 | -0.744 | .457 |
| FU12*Persisting High Risk | 0.647 | 0.458 | 611 | 1.414 | .158 | -0.601 | 1.404 | -0.428 | .669 |
| FU12*Onset | 0.498 | 0.565 | 611 | 0.881 | .378 | -2.851 | 1.566 | -1.821 | .069 |
| FU12*Subsiding | 0.746 | 0.567 | 611 | 1.317 | .188 | 1.463 | 1.891 | 0.774 | .439 |
| **Random Effects** | | | | | | | | | |
| ID Var (SD) | 21.59 (4.646) | | | | | 106.4 (10.32) | | | |
| FU12 Var (SD) | 10.33 (3.214) | | | | | — | | | |
| **Fit Indices** | | | | | | | | | |
| AIC | 7266.1 | | | | | 587.6 | | | |
| BIC | 7363.4 | | | | | 669.8 | | | |

*Note.* Table S3 presents the results for the fixed and random effects of the CannabisQuant-Model, the TobaccoBinary-Model, as well as AIC and BIC for model fit. Baseline (BL)–ID is the random effect accounting for the random intercepts between individuals; The random effects for FU6 and FU12 are the variances and standard deviations for the random slopes over time. The interaction terms represent the moderation of the effect of time of measurement by each PLE trajectory group relative to the reference (non-HIP) group. *SE* = Standard Error; *df* = degrees of freedom; *t* = t-score for the corresponding estimate. *Asterisks* indicate statistical significance. Significant *p*-values and estimates are printed in bold
